# Coronary artery calcification in the Moscow population (based on the Moscow Experiment on the use of computer vision in medical imaging)

**DOI:** 10.1101/2023.04.12.23288344

**Authors:** Yuriy A. Vasilev, Inna V. Goncharova, Anton V. Vladzymyrskyy, Igor M. Shulkin, Kirill M. Arzamasov

**Affiliations:** State Budget-Funded Health Care Institution of the City of Moscow “Research and Practical Clinical Center for Diagnostics and Telemedicine Technologies of the Moscow Health, Care Department”, 24/1 Petrovka St., Moscow, 127051, the Russian Federation

**Keywords:** coronary artery calcium, cardiovascular disease, artificial intelligence, computed tomography, opportunistic screening

## Abstract

**Introduction:** The challenges related to the prevention, diagnosis, and treatment of cardiovascular diseases remain acute. One of the measures to prevent these conditions from occurring is early detection of risk factors, one of which is coronary calcium. The latest achievements in computer vision made it possible to conduct opportunistic screening for coronary calcium.

**Objective:** To study the prevalence of coronary artery calcification (CAC) as a risk factor for cardiovascular diseases in the Moscow population based on automated analysis of imaging findings.

**Materials and methods:** A retrospective descriptive epidemiology study was carried out. Chest CT studies of 165,234 patients were analyzed. AI services carried out the automated analysis to detect CAC and to calculate the CAC score (CACS).

**Results:** Coronary calcium was detected in 61.4% of the participants. The proportion of diagnosed men was 68.9% (of all men), women – 55.7% (of all women) (p<0.001). The CAC score ranged between 1 to 60,306; the mean value was 558.2. The mean CACS increase for the entire population was 170.75; the mean growth was 168.13% the mean growth rate was 68.13%. 47.6% of men and 36.5% of women had clinically significant CACS (p<0.001). Most participants with clinically significant CAC belonged to Elderly and Senile age groups (42.0% each).

**Conclusions:** The prevalence of coronary calcium in the Moscow population was 8.03 per 1000 people. CAC (including clinically significant) was significantly more common in men. The mean CACS was significantly higher compared to the female population across most age groups. There is a continuous increase in the mean CACS with age.

## Introduction

Prevention, diagnosis, and treatment of cardiovascular diseases (CVD) stay relevant in terms of health, socio-economic and demographic agenda. Extensive scientific literature offers insights into the theory and practice of clinical prevention, the identification and alleviation of various risk factors, with ample data on the synergistic effect of both factors accumulated over the recent years [1,2]. Scholars vigorously explore the real-world evidence related to the known CVD risk factors. Given the extensive adoption of new imaging modalities, the evaluation of imaging studies for risk factors is becoming ever more important [3-5]. These, above all, include coronary artery calcium (calcium plaque build-ups found in the walls of the coronary arteries).

The bulk of research on CAC has focused on its radiographic features, diagnostic and prognostic aspects, correlations between the CAC severity and other conditions, pathologies, and outcomes.

In these circumstances, there is virtually no knowledge about the CAC incidence and relevant numerical data. Both in Russia and the rest of the world, the data on the CAC prevalence within a population are extremely limited. Available articles provide evidence only for certain gender and age groups, people at higher risk or those suffering from chronic non-communicable diseases (mostly diabetes mellitus), and certain population groups (i.e., workers occupied in certain trades) [6-9].

This situation is quite natural. There are no reports about the studies designed to detect CAC during screening programs or projects. At best, the detection of this risk factor is seen as a by-product of limited selective screening programs [10-12].

The population screening check-ups carried out in the Russian Federation under the current legislation, utilize no special techniques for detecting coronary calcification.

Chest CT remains one of the most widespread imaging modalities, both in outpatient and inpatient settings [13]. While this study is aimed to tackle various clinical and diagnostic challenges, it also opens a way to identify risk factors and early signs of high-profile diseases by means of so-called opportunistic screening. There is no doubt that making radiographers responsible for preventive check-ups will markedly prolong the reading time and consume extra resources. In addition, getting these specialists involved in routine screening may be completely unacceptable in the context of their main clinical responsibilities.

Previously, both we and other authors demonstrated the feasibility of carrying out opportunistic screening through automated analysis of diagnostic images. Such background analysis, which neither hinders nor slows down their main job, can be done using software based on artificial intelligence technologies (AI) [14-16].

In 2020, the Government of Moscow sponsored the world’s largest prospective clinical study of the feasibility and adequacy of AI in radiology – the “Experiment on the Use of Innovative Technologies in the Field of Computer Vision for the Analysis of Medical Images and Further Use in the Healthcare System of Moscow” (mosmed.ai) (hereinafter – the Moscow Experiment). We developed procedures for step-by-step evaluation of diagnostic accuracy and reliability, carried out technical and clinical monitoring, and assessed the AI impact on the safety, quality, and cost-effectiveness of medical care. This study design was based on the AI (so-called AI services) evaluation of radiological examinations obtained with various imaging modalities. At the time of writing, the Moscow Experiment had engaged over 70 AI services, which had analyzed 8.9 million imaging studies from more than 150 medical facilities in Moscow (in 2022, the Experiment was expanded to the medical facilities of the Yamalo-Nenets Autonomous Okrug).

The Moscow Experiment accumulated AI readings of chest CT scans aimed to identify various risk factors for chronic non-communicable diseases.

### Objective

To study the prevalence of coronary artery calcification (CAC) as a risk factor for cardiovascular diseases in the Moscow population based on an automated analysis of imaging findings.

## Materials and methods

The study was carried out as part of the “Experiment on the Use of Innovative Technologies in the Field of Computer Vision for the Analysis of Medical Images and Further Use in the Healthcare System of Moscow” which is being conducted since 2020 and supported by the Moscow Government (mosmed.ai) [16].

Design: a retrospective descriptive epidemiology study.

The analysis of the Moscow population data covered the period between July 2021 and December 2022.

This study is based on data from 165,234 participants. The study included participants with available data on at least one study variable. The analysis utilized a breakdown by sex and age. The age groups were built using the classification of the World Health Organization: Young – 18-44 years; Middle – 45-59 years; Elderly – 60-74 years; Senile – 75-89 years; Long-livers – 90 years or more.

The primary data were taken from the healthcare information systems of a constituent entity of the Russian Federation – the Unified Radiological Information Service of the Unified Medical Information and Analytical System of Moscow (ERIS EMIAS).

Participants were referred to chest CT by their attending physicians; the imaging procedures were performed by X-ray technicians in Moscow public inpatient or outpatient medical facilities. The Chest CT studies stored in ERIS EMIAS were sent to the software service based on artificial intelligence technologies (hereinafter – AI service) as per the protocol of the Moscow Experiment [16]. After that, both the initial studies and automated readings were made available to human radiologists for double reading, interpretation, and report-making.

The Moscow Experiment involved automated analysis of chest CT scans using AI services, two of which were designed to identify the CVD risk factors, i.e., the coronary calcium: “CVL-Chest CT Coronary calcium” (CVisionLab), “Agatston-IRA” (Intelligent Radiology Assistance Laboratories (IRA Labs)).

The applicability of the generated results for the epidemiological study is evidenced by the high diagnostic accuracy of the AI-based software. The corresponding AUC values are determined in a prospective manner through the technical and clinical monitoring carried out as part of the Moscow Experiment (Table 1). The corresponding methodologies have been published earlier [16].

**Table 1.** Diagnostic accuracy variable – AUC value for the AI services which were used to detect CAC and calculate CACS.

| Software based on artificial intelligence technologies | Area under the curve | 95% confidence interval |
| --- | --- | --- |
| CVL-Chest CT Coronary calcium | 0.83 | 0.76-0.9 |
| Agatston-IRA | 0.99 | 0.96-1.0 |

The above AI services differ in terms of functionality since at the early stages the Moscow Experiment expected only a binary assessment of the risk factors. Later, the requirements were expanded to the automated morphometric analysis of the CAC composition (measurement of CAC score and volume).

The AI services automatically evaluated chest CT studies for the following parameters: the presence of the expected risk factor (binary assessment: yes/no); the Agatston score (the score >=300 was considered clinically significant [17]).

The results of the chest CT analysis performed by the AI services are stored in ERIS EMIAS. This allowed us to analyze the features and structure of the prevalence of the CVD risk factors (i.e., coronary calcium) for the population.

The analysis and processing of the data were based on the following **methods**:

1. Statistical analysis. Descriptive statistics were used to present the following data: number of non-missing values (N), minimum value (Min), maximum value (Max), arithmetic mean (M), standard deviation (SD), 95% confidence interval (CI) for the mean, median (Me), the first and the third quartiles (Q1, Q3). A comparison of categorical data for the groups was carried out using the χ2 test. Numerical data were calculated using the analysis of variance (ANOVA). Detection of statistically significant differences was followed by the post-hoc paired t-test adjusted for the multiple comparison based on Tukey’s test. The study utilized a significance level of 0.05 (two-sided). Additionally, logistic regression models were built. A dependent variable represented the presence or absence of the risk factor for each participant. Sex, age, and age square for the non-linear relationship with age, were used as the model variables. For each variable, we estimated the odds ratio (OR) for the risk factor and 95% CI for OR. Statistical processing was performed using the Stata14® software.
2. Building and analyzing the interval time series.
3. Determining the prevalence rate. It was calculated as a ratio of the number of cases to the mid-year population, multiplied by 1000. The mid-year population was calculated as a mean of the mid-year population for 2021 – 12,645,258 people (according to public data from the Federal State Statistics Service).

Terminology clarification. In this paper, we refer to the “presence of coronary calcium”, which means the presence of coronary calcium seen through a chest CT image. Our study neither required nor utilized other methods to verify the presence and features of the risk factors.

## Results

During the specified period, the Moscow public outpatient facilities provided access to 739,140 chest CT studies; these studies were ordered by attending physicians to address various problems. Of this number, 91.3% (674,943) of the chest CT studies were analyzed by the AI services, including 165,234 (22.4% of all studies) were analyzed to identify the target risk factor.

This paper includes the studies of 165,234 patients with available binary data on CAC, of which 45,065 had a calculated CAC score (hereinafter – CACS).

CT scan results of 165,234 participants were analyzed by AI-based software to detect and carry out morphometric analysis of CAC.

The CAC risk factor was observed in 61.4% (101,528) of the examined participants (mean age 63.5+16.1, mode – 75, median – 65). The prevalence^1^ of the CAC risk factor in the Moscow population was 8.03 per 1000 people.

The proportion of diagnosed men was 68.9% (49,379 of all men), women – 55.7% (52,149 of all women). The risk factor was observed significantly more frequently in men (z-score 95.0, p<0.001).

Table 2 shows the age distribution of participants with diagnosed CAC.

**Table 2.** Distribution of participants with CAC, by age.

| Age group, years | Males | Females | Total |
| --- | --- | --- | --- |
| 18-44 | 2,373/11,759 (20.2%) | 690/10,446 (6.6%) | 3,063/22,205 (13.8%) |
| 45-59 | 10,405/17,376 (59.9%) | 4714/18,238 (25.9%) | 15,119/35,614 (42.5%) |
| 60-74 | 23,946/28,736 (83.3%) | 22,185/36,441 (60.9%) | 46,131/65,177 (70.8%) |
| 75-89 | 11,687/12,754 (91.6%) | 21,625/25,274 (85.6%) | 33,312/38,028 (87.6%) |
| 90 and older | 968/1,010 (95.8%) | 2,935/3,200 (91.7%) | 3,903/4,210 (92.7%) |
| **Total** | **49,379/71,635 (68.9%)** | **52,149/93,599 (55.7%)** | **101,528/165,234 (61.4%)** |
| **p** | **<0.001** | **<0.001** | **<0.001** |

Most participants with CAC observed on chest CT belong to the Elderly (45.0%) and Senile (33.0%) age groups. The least number of participants with the findings of interest belong to Young (3.0%).

In general, there is a direct relationship between age and the presence of CAC: 13.8% – Young; 42.5% – Middle; 70.8% – Elderly; 87.6% – Senile; 92.7% – Long-livers. A similar trend was observed separately in male and female cohorts.

In each age group, CAC was observed statistically more often in men than in women. The differences were especially pronounced in Middle (10,405 (59.9%) vs. 4,714 (25.9%), Chi-square = 4,200, p<0.001) and Elderly (23,946 (83.3%) vs. 22,185 (60.9%), Chi-square=3,900, p<0.001).

To identify the parameters influencing the risk of coronary calcium we used logistic regression. It was found that OR for CAC in men vs. women of the same age was 3.564 (95% CI 3.472; 3.659; z-score – 95.0, p<0.001). Adding 5 years to the participant’s age increases the probability of having this risk factor by 1.616 times (95% CI 1.607; 1.624; z-score 185.0, p<0.001).

Automated morphometric analysis of the CAC composition and the Agatston score was available for 45,065 participants; the results are shown in Table 3.

**Table 3.** Automated morphometric analysis of the CAC composition (CACS measurement)

| Gender | Parameter | Age group, years |
| --- | --- | --- |

<sup>1</sup> Prevalence – the ratio of the number of cases to the mean annual population \* 1000 Mean for 2021 – 12,645,258 (source: Mosgorstat).

|  |  | **18-44** | **45-59** | **60-74** | **75-89** | **90 and older** | **Total** |
| --- | --- | --- | --- | --- | --- | --- | --- |
| Total | N | 2,656 | 7,347 | 19,195 | 14,202 | 1,665 | 45,065 |
|  | Mean | 97.7 | 304.3 | 567.5 | 737.1 | 780.7 | 558.2 |
|  | SD | 292.1 | 700.6 | 996.8 | 1,210.5 | 954.1 | 1,019.1 |
|  | 95% CI | (86.6; 108.8) | (288.3; 320.3) | (553.4; 581.6) | (717.2; 757.0) | (734.9; 826.6) | (548.8; 567.6) |
|  | Min | 1 | 1 | 1 | 1 | 1 | 1 |
|  | Max | 3,550 | 23,926 | 24,820 | 60,306 | 13,008 | 60,306 |
|  | Med | 8 | 65 | 198 | 381 | 462 | 201 |
|  | Q1 | 1 | 9 | 40 | 111 | 158 | 35 |
|  | Q3 | 55 | 299 | 689 | 965 | 1070 | 688 |
|  | F(ANOVA) | 392.4 |  |  |  |  |  |
|  | P | <0.0001 |  |  |  |  |  |
| Males | N | 2,047 | 5,233 | 10,212 | 5,107 | 410 | 23,009 |
|  | Mean | 97.7 | 370.6 | 767.2 | 1,023.2 | 1,082.2 | 679.9 |
|  | SD | 288.3 | 792.9 | 1,164.6 | 1,594.6 | 1,013.2 | 1,191.3 |
|  | 95% CI | (85.2; 110.2) | (349.1; 392.0) | (744.6; 789.8) | (979.4; 1,066.9) | (983.8; 1,180.5) | (664.5; 695.2) |
|  | Min | 1 | 1 | 1 | 1 | 2 | 1 |
|  | Max | 3,550 | 23,926 | 24,820 | 60,306 | 5,645 | 60,306 |
|  | Med | 9 | 95 | 355 | 608 | 771.5 | 266 |
|  | Q1 | 1 | 15 | 86 | 206 | 286 | 45 |
|  | Q3 | 59 | 387 | 1,008 | 1,373 | 1,646 | 878 |
|  | F(ANOVA) | 363.4 |  |  |  |  |  |
|  | P | <0.0001 |  |  |  |  |  |
| Females | N | 609 | 2,114 | 8,983 | 9,095 | 1,255 | 22,056 |
|  | Mean | 97.7 | 140.3 | 340.4 | 576.4 | 682.2 | 431.3 |
|  | SD | 304.9 | 334.6 | 696.1 | 888.1 | 913.1 | 781.1 |
|  | 95% CI | (73.4; 121.9) | (126.1; 154.6) | (326.0; 354.8) | (558.2; 594.7) | (631.7; 732.8) | (421.0; 441.6) |
|  | Min | 1 | 1 | 1 | 1 | 1 | 1 |
|  | Max | 3,305 | 3,621 | 14,851 | 20,866 | 13,008 | 20,866 |
|  | Med | 4 | 25 | 102 | 288 | 401 | 155 |
|  | Q1 | 1 | 3 | 20 | 82 | 129 | 29 |
|  | Q3 | 43 | 117 | 360 | 741 | 928 | 521 |
|  | F(ANOVA) | 253.5 |  |  |  |  |  |
|  | P | <0.0001 |  |  |  |  |  |

Note: N – total number of studies in the sample, Mean – arithmetic mean, SD – standard deviation, Min – minimum value in the sample, Max – maximum value in the sample, Med – median, Q1, Q3 – values of the first and third quartiles.

CACS ranged between 1 to 60,306; the mean value was 558.2 (95% CI 548,8; 567,6).

The means varied quite significantly across different age groups and had a tendency for linear growth. The mean CAC in Young was 97.7 (95% CI 86.6; 108.8); a jump to 304.3 (95% CI 288.3; 320.3) was observed in the participants aged 45-59 years. In the older population, the increase in CACS spread more evenly: Elderly – 567.5 (95% CI 553.4; 581.6); Senile – 737.1 (95% CI 717.2; 757.0); Long-livers – 780.7 (95% CI 734.9; 826.6).

The group of 75-89 years old participants is featured by the highest CACS (up to 60,306) and the highest variability (standard deviation was 1210.5, against 292.1-996.8 in other groups).

In men, CACS varied in the above range, while the mean was 679.9 (95% CI 664.5; 695.2). The score followed a similar trend and increased with age. We observed an almost threefold increase in the mean score in Young: from 97.7 (95% CI 85.2; 110.2) to 370.6 (95% CI 349.1; 392.0) on average. Then a smoother but significant increase of around 300 was seen in the next two age groups. Only when comparing Senile and Long-livers, the difference in the means becomes negligible (1023.2 (95% CI 979.4; 1066.9) vs. 1082.2 (95% CI 983.8; 1180.5), respectively).

In women CACS ranged between 1 to 20,866; the mean was 431.3 (95% CI 421.0; 441.6). Females follow the same general trend of CACS getting higher with age.

However, the difference in the means between the 18-44 and 45-59-year-olds was less significant (97.7 (95% CI 73.4; 121.9) and 140.3 (95% CI 126.1; 154.6)). In the older age groups, there was an almost two-fold increase in CACS by about 150-200. The CACS increase slowed down only in Long-livers and reached 682.2 (95% CI 631.7; 732.8).

The mean CACS in men was 679.9 (95% CI 664.5; 695.2); women – 431.3 (95% CI 421.0; 441.6). The differences were statistically significant (t = 26.1, p<0.0001).

The analysis made it possible to reveal certain trends. A more detailed analysis was done using interval time series.

Interval time series for CACS were built and analyzed for the entire population, and for the male and female cohorts separately (Tables 4, 5, 6).

**Table 4.** Interval time series for mean CACS according to automated morphometric analysis (total sample)

| Age group | Level | Absolute rate of change from baseline | Increase | Growth coefficient |  | Growth, % | Growth rate, % |  |
| --- | --- | --- | --- | --- | --- | --- | --- | --- |
|  |  |  |  | change from baseline | change from previous |  | change from baseline | change from previous |
| 18-44 | 97.7 | - | - | - | - | - | - | - |
| 45-59 | 304.3 | 206.60 | 206.60 | 3.11 | 3.11 | 311.46 | 211.46 | 211.46 |
| 60-74 | 567.5 | 469.80 | 263.20 | 5.81 | 1.86 | 186.49 | 480.86 | 86.49 |
| 75-89 | 737.1 | 639.40 | 169.60 | 7.54 | 1.30 | 129.89 | 654.45 | 29.89 |
| >90 | 780.7 | 683.00 | 43.60 | 7.99 | 1.06 | 105.92 | 699.08 | 5.92 |

**Table 5.** Interval time series for mean CACS according to automated morphometric analysis (males)

| Age group | Level | Absolute rate of change from baseline | Increase | Growth coefficient |  | Growth, % | Growth rate, % |  |
| --- | --- | --- | --- | --- | --- | --- | --- | --- |
|  |  |  |  | change from baseline | change from previous |  | change from baseline | change from previous |
| 18-44 | 97.7 | - | - | - | - | - | - | - |
| 45-59 | 370.6 | 272.90 | 272.90 | 3.79 | 3.79 | 379.32 | 279.32 | 279.32 |
| 60-74 | 767.2 | 669.50 | 396.60 | 7.85 | 2.07 | 207.02 | 685.26 | 107.02 |
| 75-89 | 1023.2 | 925.50 | 256.00 | 10.47 | 1.33 | 133.37 | 947.29 | 33.37 |
| >90 | 1082.2 | 984.50 | 59.00 | 11.08 | 1.06 | 105.77 | 1,007.68 | 5.77 |

**Table 6.**
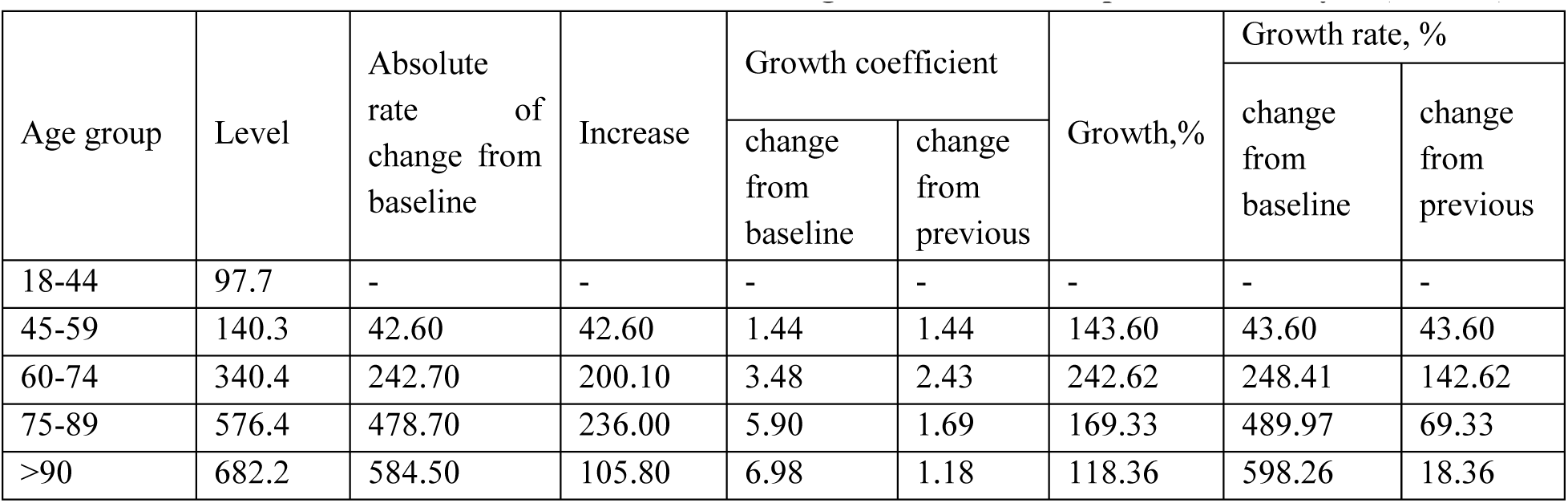
Interval time series for mean CACS according to automated morphometric analysis (females)

| Age group | Level | Absolute rate of change from baseline | Increase | Growth coefficient |  | Growth,% | Growth rate, % |  |
| --- | --- | --- | --- | --- | --- | --- | --- | --- |
|  |  |  |  | change from baseline | change from previous |  | change from baseline | change from previous |
| 18-44 | 97.7 | - | - | - | - | - | - | - |
| 45-59 | 140.3 | 42.60 | 42.60 | 1.44 | 1.44 | 143.60 | 43.60 | 43.60 |
| 60-74 | 340.4 | 242.70 | 200.10 | 3.48 | 2.43 | 242.62 | 248.41 | 142.62 |
| 75-89 | 576.4 | 478.70 | 236.00 | 5.90 | 1.69 | 169.33 | 489.97 | 69.33 |
| >90 | 682.2 | 584.50 | 105.80 | 6.98 | 1.18 | 118.36 | 598.26 | 18.36 |

The data for the general population shows a steady increase in absolute rate of change from baseline (from 206.6 in 45-59-year-old participants to 683.0 in Long-livers), which means that CACS increases with age. The highest increase was observed in Middle and Elderly – the CACS growth rate in these age groups was the most prominent, unlike the younger age groups. A pronounced jump observed in 45–59-year-olds was confirmed. Unlike the younger participants, the growth rate in this group was 311.46%; while in older groups the growth rate becomes substantially lower and even experiences a gradual decline. Thus, in the older cohorts the mean CACS becomes substantially higher (the rate of change from baseline in 45–59-year-olds was 3.11, whereas in Long-livers it went up to 7.99). At the same time, the growth tempo for this coefficient declines with age (rate of change from the previous period in 45–59-year-olds – 3.11, in older participants – 1.86, 1.3, 1.06, respectively). Long-livers demonstrate the lowest growth tempo in their mean CACS: the rate of change from the previous period drops to 5.92% (compared to 29.89% in the younger cohort), while growth falls to 43.6 (169.6 in the younger cohort).

Males demonstrate a steady increase in their mean CACS with age: The change from baseline increased from 3.79 in Middle to 11.08 in Long-livers; the rate of change from baseline went up from 279.32% to 1007.68%, respectively. However, the growth tempo slows down with age. Initially, the growth value increases from 272.9 (Middle) to 396.6 (Elderly) and then decreases to 59.0 in Long-livers. The rate of change from the previous period is the highest in people aged 45-59 (279.32%), and the lowest in Long-livers (5.77%). The growth rate is clearly declining.

The female population, in general, follows a similar trend. The rate of change from baseline rises from 1.44 to 6.98. However, women aged 45-59 demonstrate a less rapid increase in the mean CACS, compared to men of the same age group: The increase is only 42.6, compared to 272.9 in men. This figure increased in older participants; Long-livers demonstrate a decline in growth from 236.0 to 105.8, which is, however, less pronounced than that in men (from 256.0 to 59.0). The highest increase tempo is observed in participants aged 60-74 years: the maximum increase from the previous period – 2.43, growth rate – 242.62%, rate of change from the previous period – 142.62%. In Long-liver females these figures are the lowest: 1.18, 118.36%, 18.36%.

The mean CACS increase for the entire population was 170.75; the mean growth was 168.13% the mean growth rate was 68.13%. In the male cohort, these means are higher (246.13, 182.4, 82.43, respectively). On the other hand, the figures for women are lower (146.13, 162.56, 62.56). From this, it follows that the change of CACS with age is more pronounced in men.

These trends require further statistical analysis to show evidence of their significance.

The null hypothesis suggesting that the mean values across all age groups are equal was tested (Table 3). The null hypothesis was rejected both for the entire sample (f-test – 392.4, p<0.0001) and the male and female cohorts separately. Therefore, the means in at least two age groups differ from each other. The next step was a pairwise comparison of all age groups (for the entire sample and the male and female cohorts separately).

A pairwise comparison of CACS across the age groups (Table 7) made it possible to observe significant differences in all cases, except between Senile and Long-livers (p = 0.445).

**Table 7.** Results of a post-hoc pairwise comparison of CACS between age groups, adjusted for the multiple comparison based on Tukey’s test [18] (all participants)

| Comparison | Mean | 95% CI | t | P |
| --- | --- | --- | --- | --- |
| 45-49 vs 18-44 | 206.6 | 144.7; 268.5 | 9.1 | <0.001 |
| 60-74 vs 18-44 | 469.8 | 413.2; 526.3 | 22.7 | <0.001 |
| 75-89 vs 18-44 | 639.4 | 581.6; 697.2 | 30.2 | <0.001 |
| 90 and older vs 18-44 | 683.0 | 597.6; 768.4 | 21.8 | <0.001 |
| 60-74 vs 45-59 | 263.1 | 225.7; 300.6 | 19.2 | <0.001 |
| 75-89 vs 45-59 | 432.8 | 393.5; 472.0 | 30.1 | <0.001 |
| 90 and older vs 45-59 | 476.4 | 402.2; 550.6 | 17.5 | <0.001 |
| 75-89 vs 60-74 | 169.6 | 139.4; 199.9 | 15.3 | <0.001 |
| 90 and older vs 60-74 | 213.3 | 143.4; 283.1 | 8.3 | <0.001 |
| 90 and older vs 75-89 | 43.6 | -27.2; 114.4 | 1.7 | 0.445 |

Similar observations were obtained from a pairwise comparison of the male population with a breakdown by age group (Table 8). The mean values differed between all groups, except for Senile and Long-livers (p=0.858). On the contrary, in women, statistically significant differences persisted between the above groups (t-test – 4.6, p<0.001) (Table 9).

**Table 8.** Results of a post-hoc pairwise comparison of CACS between age groups, adjusted for the multiple comparison based on the Tukey’s test (males)

| Comparison | Mean | 95% CI | t | P |
| --- | --- | --- | --- | --- |
| 45-49 vs 18-44 | 272.9 | 190.7; 355.0 | 9.1 | <0.001 |
| 60-74 vs 18-44 | 669.5 | 593.1; 745.8 | 23.9 | <0.001 |
| 75-89 vs 18-44 | 925.5 | 843.0; 1007.9 | 30.6 | <0.001 |
| 90 and older vs 18-44 | 984.5 | 813.9; 1155.0 | 15.8 | <0.001 |
| 60-74 vs 45-59 | 396.6 | 343.0; 450.2 | 20.2 | <0.001 |
| 75-89 vs 45-59 | 652.6 | 590.6; 714.6 | 28.7 | <0.001 |
| 90 and older vs 45-59 | 711.6 | 550.0; 873.3 | 12.0 | <0.001 |
| 75-89 vs 60-74 | 256.0 | 202.0; 310.0 | 12.9 | <0.001 |
| 90 and older vs 60-74 | 315.0 | 156.2; 473.7 | 5.4 | <0.001 |
| 90 and older vs 75-89 | 59.0 | -102.8; 220.8 | 1.0 | 0.858 |

**Table 9.** Results of a post-hoc pairwise comparison of CACS between age groups, adjusted for the multiple comparison based on the Tukey’s test (females)

| Comparison | Mean | 95% CI | t | P |
| --- | --- | --- | --- | --- |
| 45-49 vs 18-44 | 42.7 | -53.2; 138.5 | 0.7 | 0.743 |
| 60-74 vs 18-44 | 242.8 | 155.5; 330.0 | 7.6 | <0.001 |
| 75-89 vs 18-44 | 478.8 | 391.5; 566.0 | 15.0 | <0.001 |
| 90 and older vs 18-44 | 584.6 | 481.7; 687.5 | 15.5 | <0.001 |
| 60-74 vs 45-59 | 200.1 | 149.7; 250.5 | 10.8 | <0.001 |
| 75-89 vs 45-59 | 436.1 | 385.8; 486.4 | 23.7 | <0.001 |
| 90 and older vs 45-59 | 541.9 | 467.7; 616.2 | 19.9 | <0.001 |
| 75-89 vs 60-74 | 236.0 | 205.0; 267.0 | 20.8 | <0.001 |
| 90 and older vs 60-74 | 341.8 | 279.0; 404.6 | 14.9 | <0.001 |
| 90 and older vs 75-89 | 105.8 | 43.1; 168.6 | 4.6 | <0.001 |

The detection rate of clinically significant CAC (CACS >=300) was studied (see Table 10).

**Table 10.**
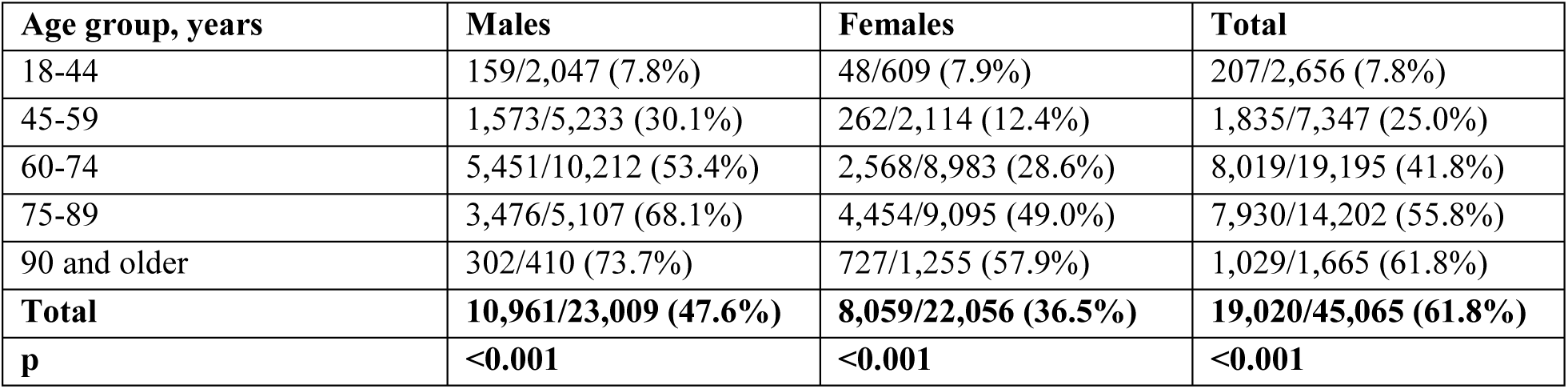
Distribution of participants with high CACS (>=300) visible in automated morphometric analysis into age groups.

| Age group, years | Males | Females | Total |
| --- | --- | --- | --- |
| 18-44 | 159/2,047 (7.8%) | 48/609 (7.9%) | 207/2,656 (7.8%) |
| 45-59 | 1,573/5,233 (30.1%) | 262/2,114 (12.4%) | 1,835/7,347 (25.0%) |
| 60-74 | 5,451/10,212 (53.4%) | 2,568/8,983 (28.6%) | 8,019/19,195 (41.8%) |
| 75-89 | 3,476/5,107 (68.1%) | 4,454/9,095 (49.0%) | 7,930/14,202 (55.8%) |
| 90 and older | 302/410 (73.7%) | 727/1,255 (57.9%) | 1,029/1,665 (61.8%) |
| **Total** | **10,961/23,009 (47.6%)** | **8,059/22,056 (36.5%)** | **19,020/45,065 (61.8%)** |
| **p** | **<0.001** | **<0.001** | **<0.001** |

A clinically significant CACS (>=300) was detected in 19,020 participants, which accounted for 61.8% of the total population with CAC.

The prevalence of the clinically significant CAC risk factor in the Moscow population was 1.51 per 1000 people.

Of all participants with CAC, 47.6% (10,961) of men and 36.5% (8,059) of women had clinically significant CACS. The difference between the male and female cohorts was statistically significant (Chi-square = 569, p<0.001).

Most participants with clinically significant CAC detected using the automated analysis, belonged to Elderly (60-74) and Senile (75-90) groups (42.0% each). The least number of participants with the findings of interest belong to Young (1.0%) and Centenarians (5.0%). The data for both sexes follow the same trend.

Of all participants with CAC, clinically significant CAC was most common to Long-livers (61.8%) and Senile (55.8%). A similar trend exists for both the male (73.7% and 68.1%, respectively) and the female cohorts (57.9% and 49.0%, respectively). Young are the least likely to develop clinically significant CACS (7.8% of all participants with this risk factor). There were no changes across the respective female and male cohorts.

With age, there was a corresponding growth in the proportion of participants with a clinically significant CACS: 7.8% – Young; 25.0% – Middle; 41.8% – Elderly; 55.8% – Senile; 61.8% – Long-livers. A similar trend was observed separately in male and female cohorts.

We found that the Young (18-44) male and female cohorts share no differences in the incidence of clinically significant CAC. However, older age groups develop more pronounced differences that acquire statical significance. The largest “gap” was observed at the age of 60-74 years: 5,451 (53.4%) vs 2,568 (28.6%), Chi-square=1,200, p<0.001).

To identify parameters influencing the risk of clinically significant CAC we used logistic regression. It was found that OR for clinically significant CAC in men vs. women of the same age was 2.792 (95% CI 2.672; 2.917; z-score – 45.9, p<0.001). Adding 5 years to the participant’s age increases the probability of having this risk factor by 1.373 times (95% CI 1.361; 1.386; z-score 68.4, p<0.001).

Interval time series for clinically significant CACS were built and analyzed for the entire population, and for the male and female cohorts separately (Tables 11, 12, 13).

**Table 11.** Interval time series for the participants with clinically significant CACS according to automated morphometric analysis (total sample)

| Age group | Level | Absolute rate of change from baseline | Increase | Growth coefficient |  | Growth, % | Growth rate, % |  |
| --- | --- | --- | --- | --- | --- | --- | --- | --- |
|  |  |  |  | change from baseline | change from previous |  | change from baseline | change from previous |
| 18-44 | 0.078 | - | - | - | - | - | - | - |
| 45-59 | 0.25 | 0.17 | 0.17 | 3.21 | 3.21 | 320.51 | 220.51 | 220.51 |
| 60-74 | 0.418 | 0.34 | 0.17 | 5.36 | 1.67 | 167.20 | 435.90 | 67.20 |
| 75-89 | 0.558 | 0.48 | 0.14 | 7.15 | 1.33 | 133.49 | 615.38 | 33.49 |
| >90 | 0.618 | 0.54 | 0.06 | 7.92 | 1.11 | 110.75 | 692.31 | 10.75 |

**Table 12.**
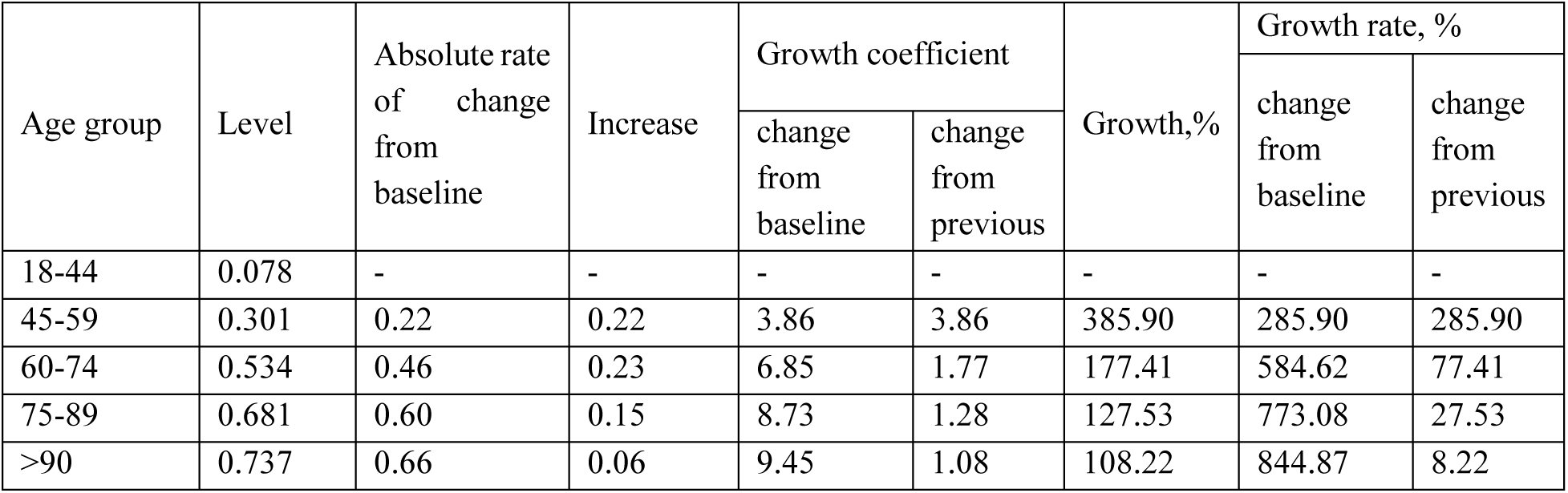
Interval time series for the participants with clinically significant CACS according to automated morphometric analysis (males)

**Table 13.**
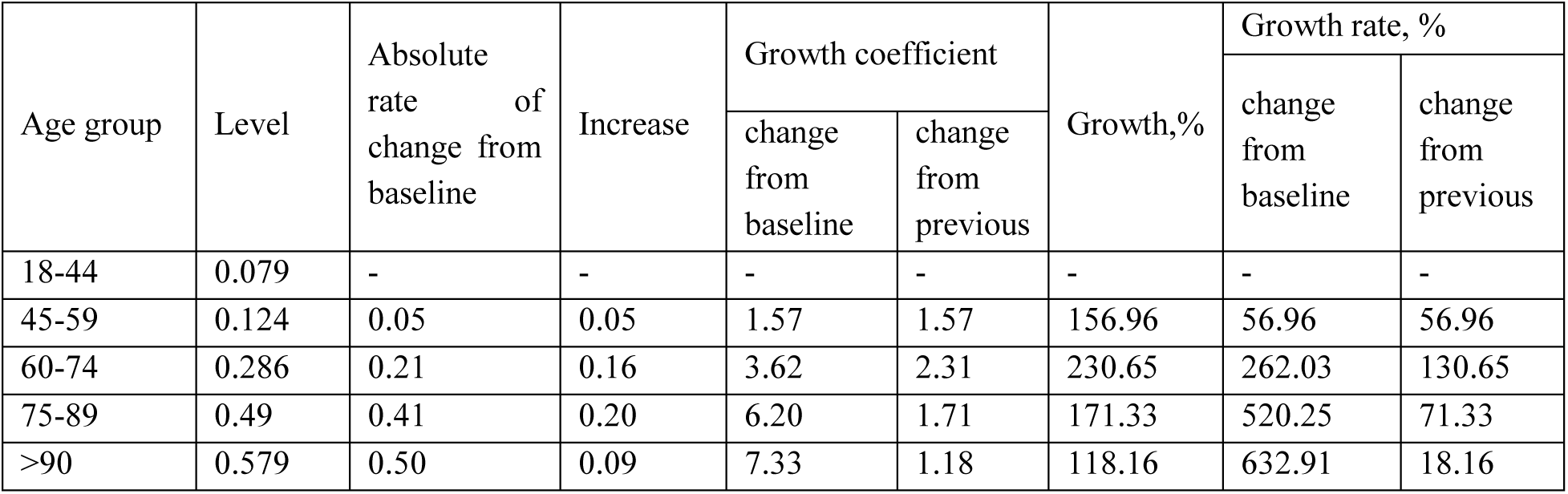
Interval time series for the participants with clinically significant CACS according to automated morphometric analysis (females)

| Age group | Level | Absolute rate of change from baseline | Increase | Growth coefficient |  | Growth, % | Growth rate, % |  |
| --- | --- | --- | --- | --- | --- | --- | --- | --- |
|  |  |  |  | change from baseline | change from previous |  | change from baseline | change from previous |
| 18-44 | 0.079 | - | - | - | - | - | - | - |
| 45-59 | 0.124 | 0.05 | 0.05 | 1.57 | 1.57 | 156.96 | 56.96 | 56.96 |
| 60-74 | 0.286 | 0.21 | 0.16 | 3.62 | 2.31 | 230.65 | 262.03 | 130.65 |
| 75-89 | 0.49 | 0.41 | 0.20 | 6.20 | 1.71 | 171.33 | 520.25 | 71.33 |
| >90 | 0.579 | 0.50 | 0.09 | 7.33 | 1.18 | 118.16 | 632.91 | 18.16 |

The data for the general population shows a steady increase in absolute rate of change from baseline (from 0.17 in 45-59-year-old participants to 0.54 in Long-livers); the same is for the change from baseline, which increased from 3.21 to 7.92 respectively. This means that with age, the proportion of participants with a significant CACS is growing steadily. However, the growth tempo slows down with age. While in Middle and Elderly the CAC increase is 0.17, in Long-livers it is only 0.06. Changes in the increase from the previous period are also apparent: It experiences a sharp two-fold drop (from 3.21 in 45–59-year-olds down to 1.67 in 60–74-year-olds) and then continues at a linear pace (down to 1.11 in Long-livers). The changes in growth and growth rates clearly illustrate the above trend: A spike in the proportion of participants with significant CACS in Middle compared to the 18–44-year-olds is followed by a linear yet less steep slope (rate of change from the previous period in 45–59-year-olds is 220.51%, while in Long-livers it is only 10.75%).

In the male cohort, the proportion of participants with significant CACS grows continuously with age (the absolute rate of change from baseline goes up from 0.22 to 0.66). In addition, Middle demonstrate the same pike in figures as the general population (growth – 385.90%, growth rate – 285.9%). Afterward, although persisting, the pace of increase notably slows down: while the growth rate in Elderly is 77.41%, in Long-livers it is only 8.22%.

For the female cohort, the situation is somewhat different. The spike is shifted and is being observed in 60–74-year-olds. Accordingly, in Middle, the proportion of women with significant CACS increases more smoothly (growth – 0.05, rate of change from the previous period – 56.96%). Elderly experience a sharp increase in figures (growth – 0.16, the rate of change from the previous period jumps to 130.65%). In the future, the proportion of participants with significant CACS continues growing (even up to the absolute growth rate of 0.5 in Long-livers), however with much less intensity (growth slows down from 171.33% in Senile to 118.61% in Long-livers).

## Discussion

The meaning of CAC (CACS) as a risk factor and a CVD predictor has been shown in a considerable number of papers and is not questioned. However, the data on its prevalence and detectability at the population level are extremely limited. The authors were indeed the first to conduct a population-based epidemiology study, made possible by such modern means of automation as artificial intelligence.

Currently, the use of artificial intelligence in medicine attracts considerable attention, as evidenced by the burst in the number of scientific papers. However, its most common applications are to automate forecasting, and diagnosing and to leverage the decision-making for health professionals [19,20]. Using the data from a population-based epidemiology study, this paper provides insight into the new way to use AI in public health studies.

Our statement is evidenced by the following publication. A review of 843 papers showed that only 2 of them addressed the use of AI in public health. The first paper is devoted to the search for relationships between the incidence, the activities of the mass media, and the public information background. The second one addresses the de-identification of patients in arrays of electronic medical records [21]. The remaining publications reflect on general concepts and ideas [22–25], which makes it safe to argue the novelty of our work.

Acknowledging that automation technologies are but a tool, we shifted the focus of our attention to the actual prevalence of risk factors for CVDs at the population level.

The only data made available to the global academic community concern the detectability of CAC in limited cohorts. For example, according to a meta-analysis of data on patients with diabetes mellitus (n=20,999), the proportion of individuals with CACS greater than 0 and greater than or equal to 100 varied between 29.3-86.0% and 22.8-65.0%, respectively [9]. Our data (61.4% and 61.8%, respectively), being a part of the above ranges, gravitate towards their upper limits.

In the group of 30–45-year-olds (n=19,725) with no signs of atherosclerotic lesions, the proportion of those with CACS of more than 0 was 16.0-26.0% in men and 7.0-10.0% in women (the lower figures were observed in Black while the higher figures were characteristic of the Caucasian participants) [8]. In our study, in Young men (18-44 years old) this figure was 20.2%, while in Young women it was 6.6%.

The scientific literature contains data on the relationships between age and gender factors and the detection of coronary calcium.

In a single-center study of a group of women in Saudi Arabia, age was found to be a predictor of particular calcium score values (an extremely limited sample of 918 women aged 55+11 years was used) [7].

In the US population, the presence and number of calcified plaques in the asymptomatic group (n=70,320) were higher in men. A steady increase in the proportion of individuals with coronary calcium along with age was registered [26].

According to a meta-analysis of 23 papers (number of patients – 20,999), it was found that in patients with diabetes mellitus, age, and male gender are risk factors for a higher CACS. A higher score value was significantly associated with an increased risk of death (for any reason) and the development of fatal or non-fatal cardiovascular disorders [9].

We did not assess the risk of acute or chronic CVD, but our demographic data allow us to confirm the findings of the above-cited meta-analysis. The probability of CAC is 3.564 times higher in men than in women, and a 5-year increase in age increases said probability by 1.616 times more (regardless of age). Thus, at the population level, the proportion of individuals with detectable CAC (incl. clinically significant CAC) does go up with age. At the same time, we found significant differences between the male and female cohorts across all age groups, except for Young (which means, that gender differences become apparent above 45 years).

The trends in the age-related changes in CAC that we have identified are in full agreement with the idea proposed by Russian scientists earlier, suggesting that CAC is an integral marker of the human biological age [27].

The results of this study allow us to offer two key recommendations. The data on real-world CAC prevalence call for revision of approaches to orchestrating population-level studies related to the screening for CVDs. At the same time, it is feasible to update the methodologies for opportunistic screening that utilize automated analysis of biomedical data. AI is applicable to the entire field of medicine and should be used as a tool for studying public health.

## Conclusions

1. This paper offers the first insight into the population-wise prevalence (i.e., the population of Moscow) of such CVD risk factor as coronary calcium: 8.03 per 1000 people. The prevalence of the clinically significant CAC risk factor (CACS >=300) is 1.51 per 1000 people.
2. Most participants with CAC seen through chest CT belong to Elderly (45.0%), while the smallest proportion was observed in Young (3.0%). Clinically significant CAC mostly occurs in Elderly (42.0%) and Senile (42.0%) across both genders.
3. This paper shares the first population data (i.e., the population of Moscow) on morphometric analysis of the CAC composition. The mean CACS in men is significantly higher compared to women across all age groups: men – 679.9 (95% CI 664.5; 695.2); women – 431.3 (95% CI 421.0; 441.6).
4. CAC (incl. clinically significant CAC) was observed significantly more often in men across all age groups, except for Young. The greatest difference was seen between Middle and Elderly, the latter having the largest proportion of participants with clinically significant CACS.
5. The mean CACS grows steadily with age, but the curve’s slope gets flatter reaching its minimum and completely losing statical significance in male Long-livers. The change of CACS with age is more pronounced in men.
6. Both genders pertaining to the 45-59 years old group, tend to develop a spike in the proportion of people with CAC (incl. clinically significant CAC).
7. OR for CAC in men vs. women of the same age was 3.564 (95% CI 3.472; 3.659; clinically significant CAC – 2.792 (95% CI 2.672; 2.917).
8. Regardless of the gender, adding 5 years to the participant’s age increases the probability of having this risk factor by 1.616 times (95% CI 1.607; 1.624), while the probability of clinically significant CAC increases by 1.373 (95% CI 1.361; 1.386).

## Supporting information

Ethics Committee approval

## Data Availability

All data produced in the present work are contained in the manuscript

## Author contributions

study concept and design: *Vasilev Yu*.*A*., *Vladzymyrskyy A*.*V*.; data collection: *Goncharova I*.*V*., *Shulkin I*.*M*., *Arzamasov K*.*M*.; analysis and interpretation of results: *Goncharova I*.*V*., *Vladzymyrskyy A*.*V*.; literature review: *Goncharova I*.*V*., *Vladzymyrskyy A*.*V*.; draft manuscript: *Goncharova I*.*V*., *Vladzymyrskyy A*.*V*., *Arzamasov K*.*M*. All authors reviewed the results and approved the final version of the manuscript.

## Compliance with ethical standards

This study is based on the results of the Experiment on the use of innovative technologies in the field of computer vision for medical images analysis and further application in the Moscow Healthcare System, approved by the Ethical Committee (extract from the protocol N 2 NEC MRO RORR dated February 20, 2020), and registered on ClinicalTrials (NCT04489992).

## Footnotes

1 Prevalence – the ratio of the number of cases to the mean annual population * 1000 Mean for 2021 – 12,645,258 (source: Mosgorstat).

